# National trends in cerebrospinal fluid biomarker testing for Alzheimer’s disease in Japan

**DOI:** 10.1101/2025.09.01.25334771

**Authors:** Masanori Kurihara, Ryoko Ihara, Kenichiro Sato, Atsushi Iwata

**Affiliations:** Department of Neurology, Tokyo Metropolitan Institute for Geriatrics and Gerontology, 35-2, Sakaecho, Itabashi-ku, Tokyo 173-0015, Japan; Integrated Research Initiative for Living Well with Dementia, Tokyo Metropolitan Institute for Geriatrics and Gerontology, 35-2, Sakaecho, Itabashi-ku, Tokyo 173-0015, Japan; Healthy Aging Innovation Center, Tokyo Metropolitan Institute for Geriatrics and Gerontology, 35-2, Sakaecho, Itabashi-ku, Tokyo 173-0015, Japan; Dementia Inclusion and Therapeutics, The University of Tokyo Hospital, 7-3-1, Hongo, Bunkyo-ku, Tokyo 113-8655, Japan

## Abstract

**Background:** Although the usefulness of cerebrospinal fluid (CSF) biomarkers for Alzheimer’s disease (AD) is well-known, their use differs widely between countries. In Japan, phospho-tau181 (p-tau181) testing for determining the cause of dementia has been covered by national health insurance since 2012 and amyloid-β42/40 ratio testing for evaluating eligibility for anti-Aβ antibodies has been covered since 2023. However, safety concerns and burdens limit their use. Here, we report national trends in CSF biomarker testing for AD in Japan.

**Methods:** We used open datasets from the national database covering nearly all health insurance claims in Japan from 2014 to 2023.

**Findings:** The annual number of health insurance claims for p-tau181 testing gradually increased from 1,358 (2014) to 3,420 (2023); 79–86% of claims were from inpatient settings throughout the study period. The number of claims was highest in the 75–79 age group and higher in women in older age. Regional variances were identified. The number of claims for CSF Aβ42/40 ratio testing in its first 3 months (December 2023 to March 2024) was 729; 56% of claims were from outpatient settings.

**Interpretation:** Although the number of health insurance claims for CSF p-tau181 testing has gradually increased, this number is low compared with the previously estimated annual diagnosis of AD. Tests are often performed in inpatient settings and regional inequity exists, whereas CSF Aβ42/40 ratio testing is slightly more often performed in outpatient settings. These data are important for understanding the current situation before the implementation of biomarker-based AD diagnosis in Japan.

**Funding:** This study was supported by the Integrated Research Initiative for Living Well with Dementia of the Tokyo Metropolitan Institute for Geriatrics and Gerontology and by the Davos Alzheimer’s Collaborative.

## Introduction

Although the usefulness of cerebrospinal fluid (CSF) biomarker testing for the diagnosis of Alzheimer’s disease (AD) is well-known,^1^ its use varies widely among countries. For example, Scandinavian and several other European countries conduct CSF biomarker testing more often than North American countries.^2^ However, its use in the Western Pacific region, including Japan, where the number of patients with dementia is increasing and over half of patients worldwide will live in the near future by 2050,^3,4^ remains to be elucidated. Considering cross-country cultural and socioeconomic differences, characterizing current trends in each country is important for the global implementation of biomarker testing for AD, including in this important region.^4,5^

Japan has several unique characteristics in the Western Pacific region. For instance, it has a high number of multidisciplinary memory clinics, the highest proportion of population aged ≥ 65 years,^4^ and wide coverage by the national health insurance system.^6^ The measurement of CSF phospho-tau181 (p-tau181) levels for determining the cause of dementia has been covered by national health insurance since 2012. Other biomarkers (e.g., amyloid-β42 [Aβ42]) have also been measured in academic hospitals using research funding.^7–10^ CSF Aβ42/40 ratio testing and amyloid positron emission tomography (PET) for evaluating eligibility for anti-Aβ antibodies have been covered by insurance since December 20, 2023. However, safety concerns, burden, and preparedness of healthcare professionals for lumbar puncture (LP) may have limited its use.

As less invasive and easier blood-based biomarkers have been approved in the United States,^11,12^ understanding national trends in CSF biomarker testing for AD in Japan is important for the wider implementation of biomarker-based AD diagnoses.^13,14^ Therefore, we investigated national trends in CSF biomarker testing for AD in Japan.

## Methods

We used open datasets from the national database (NDB), managed by the Ministry of Health, Labour and Welfare of Japan, which collects information on all health insurance claims in Japan, including but not limited to medical tests.^6,15–17^ In Japan, nearly all patients visiting hospitals and clinics use national health insurance. All health insurance claims should be collected for outpatients,^6,16^ whereas inpatient costs are either claimed on an individual test/procedure basis or using a fixed daily tariff based on the diagnosis procedure combination/per diem payment system (DPC/PDPS).^6,16^ As CSF p-tau181 is not included as a unique procedure in the DPC/PDPS, the number of inpatients receiving tests may be underestimated.

The number of claims for outpatient and inpatient CSF p-tau181 testing was collected for each fiscal year from 2014 to 2022. The proportion of inpatient testing was calculated for each year. We also collected the number of claims for “management fee for dementia differential diagnosis in specialty clinics” by certified Centers for Dementia-Related Diseases (CDRDs) to approximate the annual number of new differential diagnoses of dementia in Japan. As not all hospitals and clinics conducting differential diagnoses for dementia can claim this cost, this number is the minimum estimate of the annual number of new differential diagnoses. The numbers of claims for different sexes, age groups, and prefectures were also obtained. In NDB datasets, numbers lower than 10 were not reported for privacy reasons. Population data for each prefecture were obtained from the e-Stat Portal Site of Official Statistics of Japan^18^ to calculate the number of claims per population for each prefecture. Amyloid PET and CSF Aβ42/40 ratio measurements using Lumipulse assays have been covered by national health insurance since December 20, 2023, the same day lecanemab became available. A confirmation of brain amyloid pathology by amyloid PET or CSF Aβ42/40 ratio was required for eligibility for anti-amyloid-β antibodies. As costs for p-tau181 testing cannot be claimed in combination with Aβ42/40 ratio testing, we decided not to include the number of claims for p-tau181 testing in fiscal year 2023. Although data are currently available for only 3 months (December 2023 to March 2024 in fiscal year 2023), we also summarized the number of health insurance claims for CSF Aβ42/40 ratio.

### Role of the funding source

The funders did not have any involvement in study design, data collection, analysis, interpretation, or writing.

## Results

The annual number of health insurance claims for CSF p-tau181 testing gradually increased from 1,358 to 3,420 from 2014 to 2022 (**Figure 1A**). Throughout this period, 79–86% of claims originated from inpatient settings. The number of differential diagnoses of dementia in the CDRDs also increased from 18,430 to 28,998. The ratio between the number of CSF p-tau181 tests and diagnoses in CDRDs increased by 1·59-fold during this period (**Figure 1B**).

**Figure 1.**
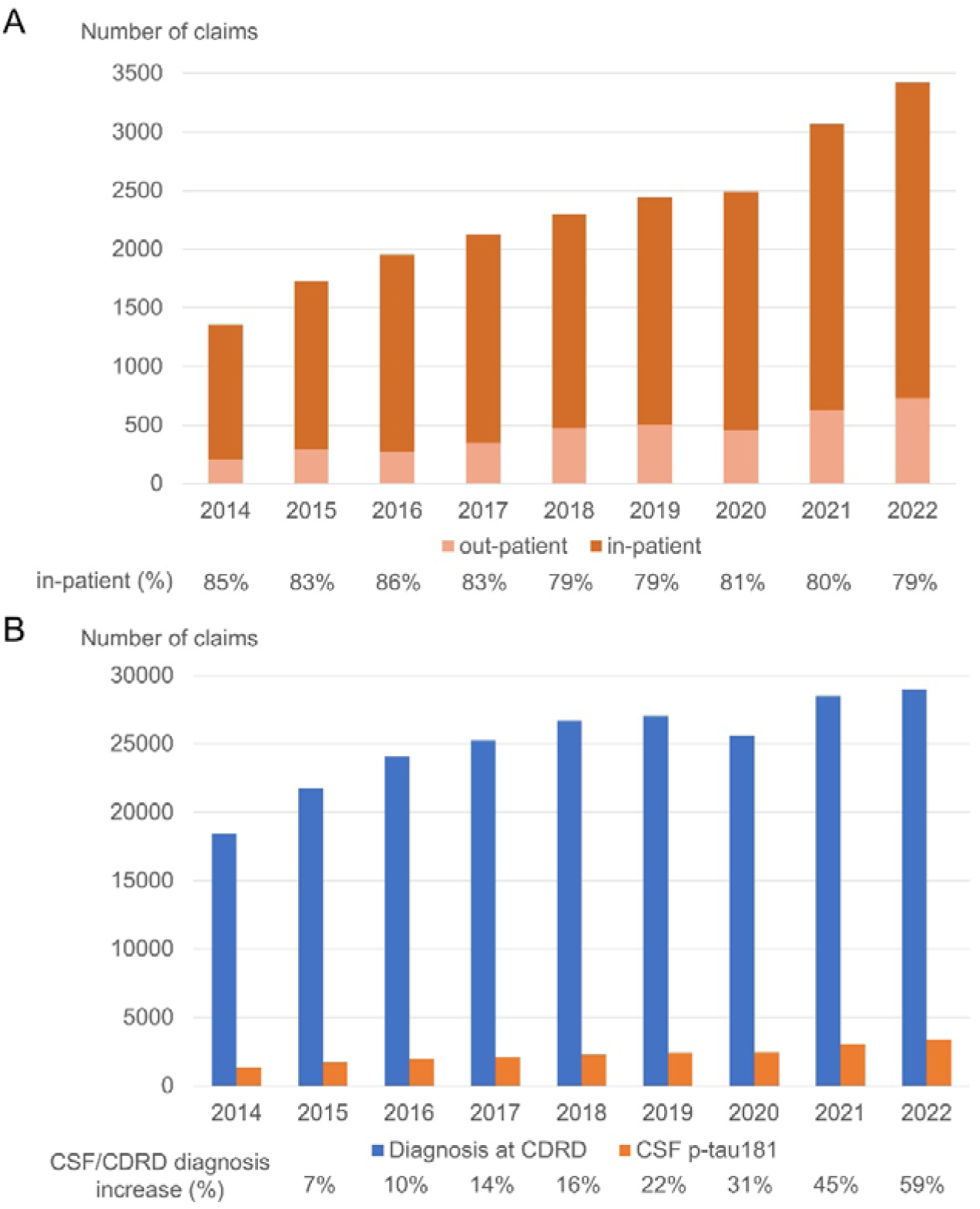
Trends in CSF p-tau181 health insurance claims in Japan. (A) Number of inpatient and outpatient claims for each fiscal year. The numbers increased during the period and the majority (≈80%) were inpatient claims. (B) Comparison with the number of differential dementia diagnosis claims from the CDRDs. The number of differential dementia diagnoses at CDRDs also increased during the period. The ratio between the number of CSF p-tau181 tests and diagnoses at CDRDs increased 1·59-fold during the period. Note that there are many hospitals and clinics conducting dementia differential diagnosis in addition to CDRDs; thus, these numbers are the minimum estimate of differential dementia diagnoses in each year. CDRD, Centers for Dementia-Related Disease; CSF, cerebrospinal fluid

The numbers of inpatient and outpatient claims were highest in the 75–79 age group and higher in women in older age (inpatient ≥ 80; outpatient ≥70) (**Figure 2**). Regional variations were observed in the number of claims and claims per population (**Figure 3**). While the highest prefecture had 44.3 annual claims per 1,000,000 people, the lowest prefectures had less than 5 annual claims per 1,000,000 people (**Figure 3**).

**Figure 2.**
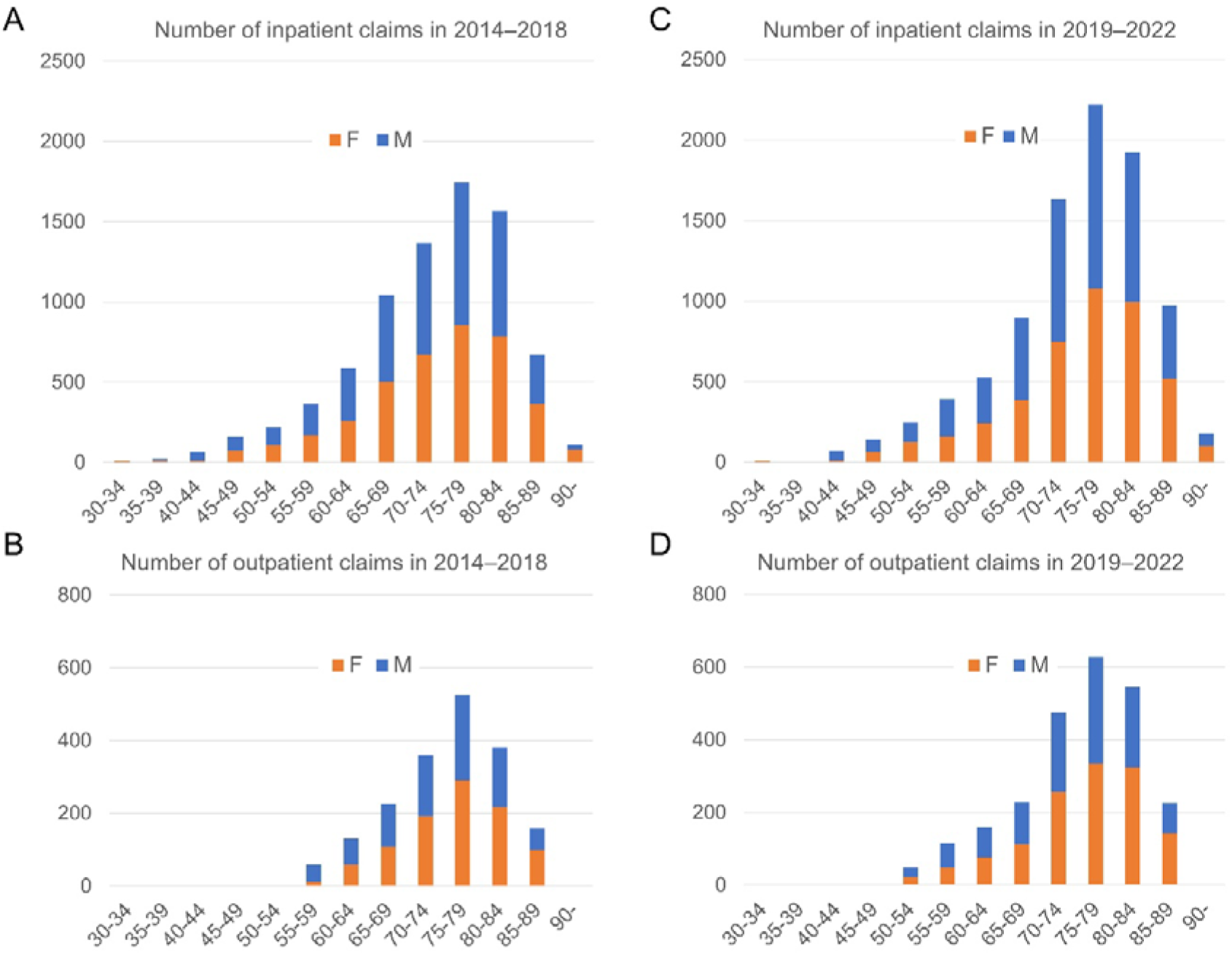
Age and sex distribution during 2014–2018 and 2019–2022. Health insurance claims for CSF p-tau181 testing in 2014–2018 (A, B) and 2019–2022 (C, D) for different age groups. The numbers of inpatient (A, C) and outpatient (B, D) claims were the highest in the 75–79 age group in both periods and were higher in women (>50%) in older age (inpatient ≥ 80; outpatient ≥70). CSF, cerebrospinal fluid

**Figure 3.**
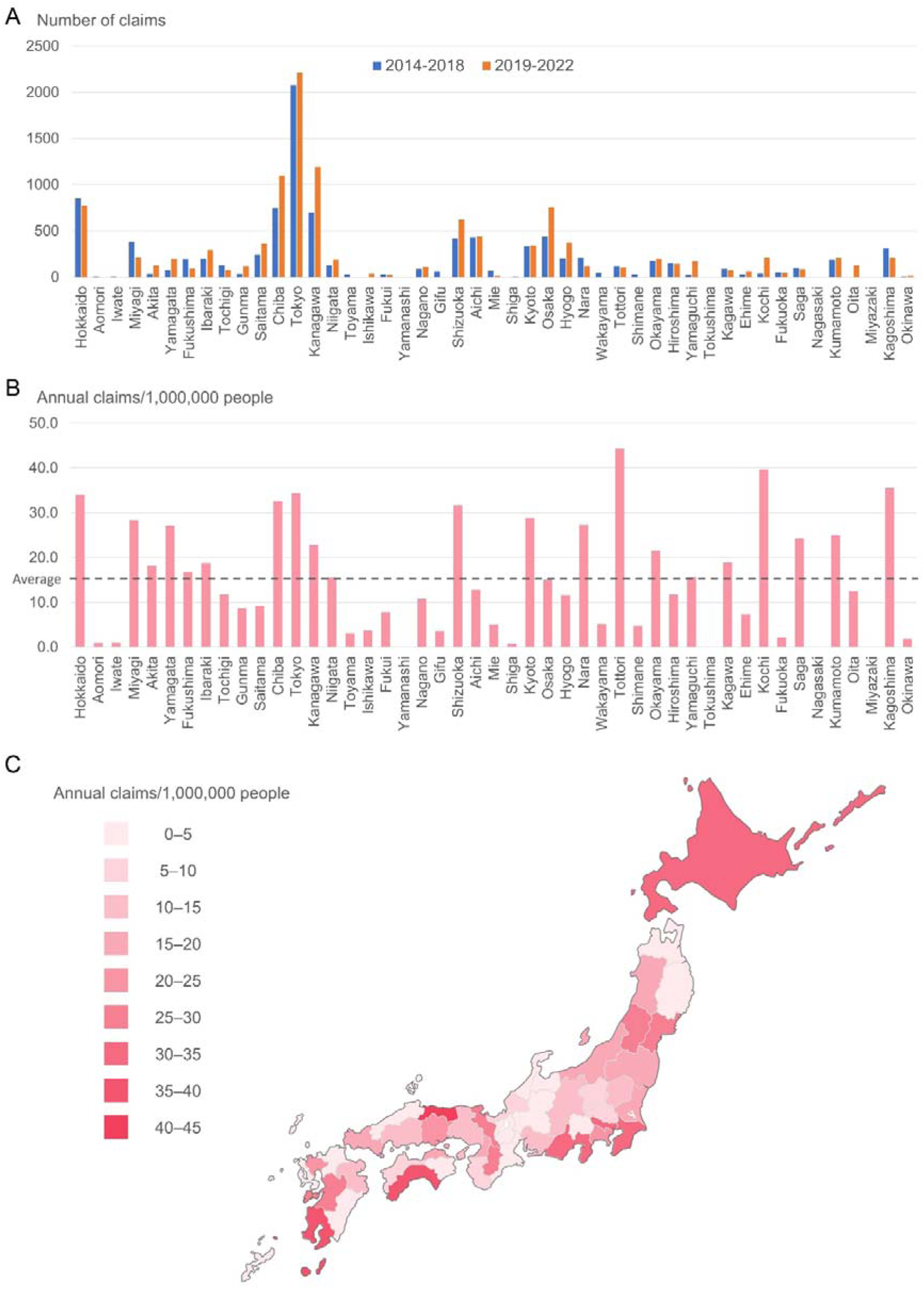
Prefectural characteristics of CSF p-tau181 health insurance claims. (A) Number of claims in each prefecture during 2014–2018 and 2019–2022. The number widely differed between prefectures, the highest being in Tokyo. The trend was similar between 2014–2018 and 2019–2022. (B) Annual claims per 1,000,000 people in each prefecture. The average annual number of claims in 2014–2022 was divided by the population of each prefecture in 2018. Even after adjustment by population, regional variances remained large between prefectures. (C) A choropleth map of the data presented in (B). CSF, cerebrospinal fluid

The numbers of amyloid PET and CSF Aβ42/40 ratio tests in fiscal year 2023 are summarized in **Table 1**. During this period, total claims for CSF Aβ42/40 ratio testing were 729 and were slightly more frequent in outpatient settings (55·7%) than in inpatient settings. As data were available for only the first 3 months, the number of claims per month were still continuing to increase (**Table 1**).

**Table 1.** Number of health insurance claims for CSF Aβ42/40 ratio testing in fiscal year 2023.

|  | Number of claims for CSF Aβ42/40 |  |  | Total | Proportion |
| --- | --- | --- | --- | --- | --- |
|  | Dec, 2023 | Feb, 2024 | Mar, 2024 |  |  |
|  | Jan, 2024 |  |  |  |  |
| Outpatient | 80 | 137 | 185 | 406 | 55.7% |
| Inpatient | 60 | 115 | 148 | 323 | 44.3% |

The numbers are from December 20, 2023 (day of coverage) to the end of the fiscal year, March 31, 2024. The total number of claims for CSF Aβ42/40 ratio testing was 729, whereas that for amyloid PET was 936 during the period. As the period was just after health insurance coverage (only for 3 months) and the number is still growing from December/January,
February, to March, these trends may change in the next fiscal year.
Aβ , amyloid-beta; CSF, cerebrospinal fluid; PET, positron emission tomography

## Discussion

We reported national trends in CSF p-tau181 testing in Japan using the NDB of health insurance claims from the fiscal years of 2014 to 2022 and summarized trends in CSF Aβ42/40 ratio testing in the first 3 months of its coverage.

Although the annual number of health insurance claims for p-tau181 testing gradually increased from 1,358 to 3,420 during the study period, these numbers were extremely low compared with the estimated annual incidence of new hospital visits. In Japan, a recent study estimated that the annual incidence of “AD and other dementia” in 2021 was 576,270.^19^ Estimates in another recent study in 2024 were 608,741 for dementia and 1,449,385 for mild cognitive impairment.^4^ Previous surveys have reported that 50–65% of patients with dementia in Japan have a clinical diagnosis of Alzheimer’s disease.^20,21^ Although the number of patients visiting specialty clinics for differential diagnosis remains unknown, the number of insurance claims in CDRDs in 2021 was 28,509,^15^ leading to an approximate estimate of 14,250–18,530 claims for AD. This is the minimum estimated number of new annual hospital visits for the differential diagnoses of AD. Possible reasons for the relatively low rate of CSF biomarker testing include safety concerns regarding LP, burdens for patients and healthcare professionals, and the lack of disease-modifying treatments during this period. The number of biomarker tests may have changed after the launch of the anti-amyloid-β antibodies lecanemab and donanemab and may change further when blood-based biomarkers become available owing to low safety concerns and burden and wide availability in most hospitals and clinics. Further research is important to estimate the demand for AD biomarkers in other countries, especially in the Western Pacific region, where research studies have been limited despite growing number of patients.^3^

During the study period, the majority (≥80%) of CSF p-tau181 tests in Japan were likely conducted in inpatient settings. Although the price estimates of LP (∼ 160 USD)^22^ and LP plus CSF biomarker testing (∼ 470–1000 USD)^11,22,23^ in the United States are high, the cost of LP in Japan is fixed by the national healthcare system at only 18 USD (exchange rate: USD 1 = JPY 147). In Japan, only physicians are permitted to perform LP. As one physician and at least one healthcare professional (e.g., nurse) are occupied for nearly 30 min, physicians may think that the procedure is not financially worthwhile.^2^ Moreover, limited time is available in most outpatient clinics in Japan owing to the fixed time schedule. In addition to the need for careful follow-up for complications, these factors may be associated with the high proportion of CSF p-tau181 tests conducted in inpatient settings. At our hospital, nearly all CSF biomarker tests for AD are conducted by residents and fellows in inpatient settings. As hospital admission not only has disadvantages for patients (e.g. cost, time, and risk of confusion/delirium) but also increases the total healthcare cost paid by insurance, reconsideration of the technical costs for LP to a higher price may be warranted.

In this study, the number of claims for CSF p-tau181 testing was highest in the 75–79 age group and was higher in women in older age (inpatient ≥ 80; outpatient ≥70). Similarly, previous studies have reported that the prevalence and incidence of dementia are higher in older patients (at least until 90).^19,24–26^ In the Japanese guidelines, CSF biomarker testing may be considered when cognitive decline is observed and the physician believes that determining or excluding a cause of dementia based on underlying pathology is clinically beneficial.^27^ Although tests are not limited to early-onset cases (<65 years), they may be less frequently conducted in older patients aged >80 years. Female dominance has also been reported in previous older age-matched studies investigating the incidence of AD dementia.^25,26^

Regional variance in CSF p-tau181 testing was identified. Tokyo had the highest number of claims, followed by Kanagawa, Chiba, Hokkaido, and Osaka (**Figure 3**). The difference remained large even after adjusting for population size, which suggests regional inequity. More in-depth studies, including data analyses and questionnaires, are necessary to elucidate the reasons for the observed differences.

We also summarized the number of claims in the first 3 months of CSF Aβ42/40 ratio testing coverage. There were already 729 claims and the number per month continued to grow from December/January, February, to March (**Table 1**). These claims were slightly more frequent in the outpatient clinics (55·7%). Differences in the proportion of inpatient versus outpatient claims between p-tau181 (2014–2022) and Aβ42/40 (2023) testing could be due to several reasons, including differences in the preparedness of hospitals/doctors and the higher price of Aβ42/40 ratio testing, which may lead to reduced profit if measured in inpatient settings owing to the fixed admission fee per day by the DPC/PDPS. However, these early data may change in the next fiscal year and should be examined over a longer time period in the future. The strength of this study lies in its use of the NDB covering nearly all health insurance claims in Japan, as national data should yield better estimates than data obtained from a limited population. However, this study also has some limitations. First, patients relying only on publicly funded healthcare are not included in the NDB dataset from 2014 to 2022; However, as this population comprises only a small proportion of patients in Japan, the effect should be minimal.^28^ Indeed, from the 2023 NDB open dataset, only 1.4% (10/729) of claims for CSF Aβ42/40 ratio were from patients only on publicly funded healthcare.^15^ Second, since we reported the number of health insurance claims, tests conducted without claims are not included. Measurements by research fundings, or some of the inpatient testing in hospitals based on DPC/PDPS may not be included. Third, there could be differences in the reason for conducting LPs between settings (e.g., to rule out AD in rapidly progressing dementia in inpatient settings), biasing these data toward a higher proportion of inpatient CSF p-tau181 tests. Forth, numbers lower than 10 are not reported in the NDB open dataset for privacy reasons; thus, age groups or prefectures with low numbers could be underestimated.

In conclusion, although the number of CSF p-tau181 tests has increased over the study period, the numbers are low compared with the estimated annual diagnosis of AD. Additionally, tests are often performed in inpatient settings and regional inequity may exist. These data are important for understanding the current situation before fully implementing biomarker-based AD diagnosis in Japan.

## Data Availability Statement

All data supporting the findings of this study are included in the manuscript or are available in the NDB^15^ (in Japanese) or on the e-Stat^18^ website.

## Acknowledgments

We thank the NDB run by the Minister of Health, Labour and Welfare and the e-Stat run by the Japanese Government for the data. This study was supported by the Integrated Research Initiative for Living Well with Dementia of the Tokyo Metropolitan Institute for Geriatrics and Gerontology and by the Davos Alzheimer’s Collaborative.

## Declaration of interests

MK received lecture fees from Eisai, Eli Lilly, and John Wiley & Sons; patent assignment fees from FUJIREBIO; and research support from Nihon Medi-Physics.

RI received advisory fees from Eli Lilly and lecture fees from Eisai and Eli Lilly.

KS is affiliated to “Dementia Inclusion and Therapeutics,” an endorsed course funded by Effissimo Capital Management Pte Ltd. KS has no other conflicts of interest to disclose. AI has received advisory fees from Eisai and Eli Lilly; stock ownership/capital gains from Eisai; lecture fees from Eisai, Eli Lilly, John Wiley & Sons, FUJIREBIO, HU Frontier, and Sysmex; and grants for commissioned/joint research from Eisai, Eli Lilly, FUJIREBIO, and Sysmex.

## Author contributions

Masanori Kurihara : Conceptualisation, Data curation, Formal analysis, Data interpretation, Visualisation, Writing – original draft.

Ryoko Ihara : Data interpretation, Writing – review & editing

Kenichiro Sato : Data interpretation,Validation, Writing – review & editing

Atsushi Iwata : Data interpretation,Funding acquisition, Writing – review & editing

## Notes

### Competing Interest Statement

MK has received lecture fees from Eisai, Eli Lilly, and John Wiley & Sons; patent assignment fee from FUJIREBIO; and research support from Nihon Medi-Physics.
RI has received advisory fees from Eli Lilly; and lecture fees from Eisai and Eli Lilly.
KS's affiliation Dementia Inclusion and Therapeutics is an endorsed course funded by Effissimo Capital Management Pte Ltd. KS has nothing else to disclose.
AI has received advisory fee from Eisai and Eli Lilly; stock ownership/capital gain from Eisai; lecture fees from Eisai, Eli Lilly, John Wiley & Sons, FUJIREBIO, HU Frontier, and Sysmex; and grants for commissioned/joint research from Eisai, Eli Lilly, FUJIREBIO, and Sysmex.

### Funding Statement

This study was supported by Integrated Research Initiative for Living Well with Dementia of Tokyo Metropolitan Institute for Geriatrics and Gerontology, and by Davos Alzheimer's Collaborative.

### Author Declarations

national database (NDB) run by the Ministry of Health, Labour and Welfare of Japan (https://www.mhlw.go.jp/stf/seisakunitsuite/bunya/0000177182.html)(in Japanese) e-Stat website (https://www.e-stat.go.jp/)

